# High prevalence of anxiety and depression among patients hospitalized with acute febrile illness in Sri Lanka

**DOI:** 10.1101/2024.02.16.24302924

**Authors:** Champica K. Bodinayake, Ajith De. Silva Nagahawatta, Wasana S. Ranaweera Arachchige, Senuri C Sahabandu, Aruna H. Pushpakumara, Piumi K. Wijayaratna, Bhagya N. Jayasekera, L. Gayani Tillekeratne

**Author notes:** Corresponding author : Champica K. Bodinayake, Faculty of Medicine, University of Ruhuna, PO Box 70, Galle, 80000 Sri Lanka.

## Abstract

**Introduction:** Acute febrile illnesses (AFI) such as dengue, leptospirosis, and influenza result in large numbers of hospitalization in the tropics. These illnesses may negatively impact patients’ mental health. We evaluated the prevalence of anxiety and depression and risk factors associated with these conditions in patients hospitalized with AFI.

**Methodology:** From July-October 2017, we conducted a cross-sectional study of consecutive patients ≥12 years admitted with AFI to a tertiary care hospital in Sri Lanka. We administered the Hospital Anxiety and Depression Scale (HADS) validated in the local language of Sinhala to identify the presence and severity of anxiety and depression. We identified the prevalence of anxiety and depression and determined the association with sociodemographic and clinical characteristics using Chi square and t-tests and multivariabele logistic regression.

**Results:** Of 193 enrolled, 44.6% were male. AFI included dengue (62.7%), viral fever (10.4%), urinary tract infection (6.2%), leptospirosis (4.7%), pneumonia (3.1%) and typhus (0.5%). Overall, 41.4% patients had anxiety and 42.5% had depression. On multivariable analysis, anxiety was associated with awareness of diagnosis (OR=4.3, CI 1.3-13.76, p=.03), lower satisfaction score (OR=0.73, CI 0.58-0.92, p=.008) and lower income (OR=1.92, CI 1.03-3.57, p=.03). Depression was associated with anorexia (OR=2.8, CI 1.33-5.87, p=.006) and diarrhea (OR=2.29, CI 1.17-4.47, p=.01).

**Conclusions:** Patients hospitalized with AFI had a high prevalence of anxiety and depression. Gastrointestinal symptoms, lower socioeconomic status, and awareness of illness diagnosis were all associated with anxiety. Our findings may be helpful in developing interventions to counteract anxiety and depression among patients hospitalized with AFI.

## Introduction

Acute febrile illness (AFI) is a common cause of hospitalization in the tropics and sub-tropics (1). Worldwide, few studies report on the mental health of patients hospitalized with AFI (2, 3). Published studies indicate a high prevalence of anxiety and depression among patients with dengue and other AFI caused by ebola, influenza and SARS-CoV-2 (4-9). Illnesses such as acute dengue, in particular, may cause psychological distress among those affected due to the potentially life-threatening nature of the disease (10). Anxiety and depression may have short and long-term ramifications in terms of disease outcome and illness recovery for both the patient and the family (11-13).

In this study, we sought to evaluate the prevalence and the severity of anxiety and depression in patients both with AFI admitted to a tertiary care center. We investigated sociodemographic, clinical and other risk factors associated with anxiety and depression to assist in the development of coping strategies and support systems to improve mental health among patients with AFI.

## Methodology

### Study Cohort

We conducted a cross-sectional study at Teaching Hospital Karapitiya (THK), Galle, the largest tertiary care center (1,800 bed) in the Southern Province of Sri Lanka. We enrolled consecutive patients ≥12 years admitted to THK with self-reported or documented fever within one week of admission, from 1^st^ of July to 30^th^ September 2017. Study doctors verified eligibility and obtained written informed consent from patients or parents for children 12-17 years of age. Ethical clearance was obtained from the Ethical Review Committee of the Faculty of Medicine, University of Ruhuna, Sri Lanka.

### Definitions

Since dengue is the most common reason for hospitalization due to AFI in our setting, we analyzed and non-dengue AFI groups separately. We defined acute dengue as patients who had a positive dengue NS1 antigen test or a positive dengue IgM antibody test (if fever >5 days). Patients who were test negative for dengue or who had an alternative clinical diagnosis confirmed or supported by other investigations were considered as non-dengue AFI.

### Study Instruments

The Hospital Anxiety and Depression Scale (HADS) developed by Zigmond and Snaith to measure both anxiety and depression among general medical patients was used in this study (14). The Sinhala translation of this tool has been validated in Sri Lanka (15). The validated Sinhala version was administered to patients by the trained study doctors. HADS has 7 items each to estimate the level of anxiety and depression, with each item scored on a scale of 0-3, giving a total score ranging from 0-21 for anxiety and depression each. A HADS score ≥8 indicated the presence of anxiety or depression and the level of anxiety or depression was graded as mild (8-11), moderate (12-14) or severe (15-21) according to the HADS instrument.

We collected information about patients’ self-perceived knowledge regarding 6 domains: the current illness diagnosis, etiology, symptoms, treatment, complications and possible preventive measures. We used a Likert scale, with each domain having a score of 0-2, totaling to a maximum of 12. Similarly, we collected information about patients’ satisfaction within the three domains of their medical care, nursing care, and facilities provided by the hospital, with each domain scored on a 0-4 scale and a maximum total of 12.

### Statistical Methods

We determined the proportion of patients with acute dengue versus non-dengue AFI and the median and interquartile range (IQR) scores on the HADS and Likert scales. We determined the association between patient’s sociodemographic and clinical characteristics with the presence of anxiety or depression. We used the Chi square test for categorical variables and the t-test for continuous variables. Independent variables with significance of p< 0.05 were entered into a multivariable logistic regression model to determine factors that were associated with anxiety and depression separately. All statistical analyses were conducted using STATA, version 11 (STATACorp, College Station, Texas).

## Results

### Sociodemographic and Clinical Characteristics in Study Cohort

Of a total of 193 enrolled patients, 86 (44.6%) were male and the median age was 35 years (IQR 25-53; Table 1). Acute dengue was confirmed in 121 (62.7%) by laboratory testing. Clinical diagnoses among the non-dengue AFI group included unspecified viral fever (20, 10.4%), urinary tract infection (12, 6.2%), leptospirosis (9, 4.7%), pneumonia (6, 3.1%) and scrub typhus (2, 0.5%). The majority of patients in this cohort (179, 92.8%) reported having family support during this acute illness. Most patients reported symptoms such as anorexia (141, 72.1%), headache (135, 70.0%), and arthralgia/ myalgia (142, 73.6%).

**Table 1.** Sociodemographic and clinical characteristics of patients with dengue and non-dengue acute febrile illness in Sri Lanka.

|  | **Total**<br>**number**<br>**(%) or**<br>**median**<br>**(IQR)**<br>**n= 193** | **Dengue number**<br>**(%) or median**<br>**(IQR)**<br>**n= 121** | **Non-dengue AFI**<br>**number (%) or median**<br>**(IQR)**<br>**n= 72** | **p-**<br>**value** |
| --- | --- | --- | --- | --- |
| **Sociodemographic characteristics** |  |  |  |  |
| Age (years) | 35 (25-53) | 32 (23- 47) | 42.5 (24.0- 63.0) | **.01** |
| Male | 86 (44.6) | 61 (50.4) | 25 (34.7) | **.03** |
| Hospital distance (km) | 13.0 (5-25) | 13.0 (5.0- 20.5) | 15.0 (5.5- 30.0) | .19 |
| Education |  |  |  |  |
| Up to GCE O/L (10<sup>th</sup> grade) | 52 (26.9) | 29 (24.0) | 23 (31.9) | .27 |
| Passed GCE O/L (10<sup>th</sup> grade) | 75 (38.9) | 46 (38.0) | 29 (40.3) |  |
| GCE A/L or above (12<sup>th</sup> grade) | 66 (34.2) | 46 (38.0) | 20 (27.9) |  |
| Occupation |  |  |  |  |
| Office/ factory/ business | 53 (27.7) | 40 (33.1) | 13 (18.1) | **.01** |
| Laborer/ agriculture | 38 (19.7) | 26 (21.5) | 12 (16.7) |  |
| Unemployed/ retired/<br>student | 102 (52.9) | 55 (45.5) | 47 (65.3) |  |
| Marital status- married | 123 (63.7) | 73 (60.3) | 50 (69.4) | .12 |
| Income per month |  |  |  |  |
| ≤25,000 Rs/ ≤162.86 USD* | 72 (37.3) | 43 (35.5) | 29 (40.3) | .54 |
| >25,000 Rs/ >162.86 USD* | 121 (62.7) | 78 (64.5) | 43 (59.7) |  |
| Family support present | 179 (92.8) | 113 (93.4) | 66 (91.7) | .66 |
| **Clinical features** |  |  |  |  |
| Days fever prior to admission | 3 (2-4) | 3 (2-4) | 3 (2-5) | **<.001** |
| Days of hospitalization at enrollment | 3 (2-4) | 3 (2-4) | 3 (2-5) | **.04** |
| Anorexia | 141 (73.1) | 89 (73.6) | 52 (72.2) | .84 |
| Headache | 135 (70.0) | 91(75.21) | 44 (61.1) | **.04** |
| Vomiting | 97 (50.3) | 60 (49.6) | 37 (51.4) | .81 |
| Diarrhea | 53 (27.5) | 37 (30.6) | 16 (22.2) | .21 |
| Arthralgia/ myalgia | 142 (73.6) | 99 (81.8) | 43 (59.7) | **<.001** |
| Cough | 44 (22.8) | 29 (24.0) | 15 (20.8) | .62 |
| Abdominal pain | 49 (25.4) | 24 (19.8) | 25 (34.7) | **.02** |
| Comorbidities<sup>+</sup> | 54 (28.0) | 22 (18.2) | 32 (44.4) | **<.001** |
| Receipt of oral medications | 188 (97.4) | 117 (96.7) | 71 (98.6) | .43 |
| Receipt of intravenous medications | 69 (35.8) | 24 (19.8) | 45 (62.5) | **<.001** |
| Receipt of intravenous fluid | 167 (86.5) | 110 (90.9) | 57 (79.2) | **.03** |
| **Patients' perceptions regarding their illness** |  |  |  |  |
| Patient awareness of diagnosis | 172 (89.1) | 115 (95.0) | 57 (79.2) | **.001** |
| Self-perceived knowledge score (out of maximal 12) | 5 (2-6) | 5 (3.5-6) | 2 (0-5) | **<.001** |
| Overall satisfaction score (out of maximal 12) | 9 (9-11) | 9 (9-11) | 9 (9-11) | 0.94 |

Abbreviations: AFI = acute febrile illness; GCE O/L = General Certificate of Education
Ordinary Level, 10<sup>th</sup> grade; GCE A/L = General Certificate of Education Advanced
Level, 12<sup>th</sup> grade. \* 1 USD = 153.51 Rs in July 2017 (22); <sup>+</sup>Diabetes mellitus,
hypertension, dyslipidemia, ischemic heart disease, asthma.

Sociodemographic features were not significantly different among dengue versus non-dengue AFI, with the exception of occupation. Patients with dengue were less likely to be unemployed or retired compared to patients with non-dengue AFI (45.5% versus 65.3%, respectively). Patients with dengue were significantly more likely to be symptomatic with headache (75.2% versus 61.1%, p=.03) and arthralgia/myalgia (81.8% versus 59.7%, p<.001) than patients with non-dengue AFI.

### Self-Reported Knowledge Score

Almost all (115, 95%) patients with dengue and a majority (57, 79%) of patients with non-dengue AFI were aware of their current illness diagnosis, as conveyed to them by their treating physicians. The median overall score for self-reported knowledge regarding their illness was 5 (IQR 2-6) in the overall study group. A significant difference between the self-reported knowledge scores was present between dengue and non-dengue AFI groups (5, IQR 3.5-6 versus 2, IQR 0-5, p<.001). The source of knowledge was reported as media by 50% and as health care personnel by 40%. Two-thirds of dengue patients (81, 66.9%) reported their source of knowledge as media, compared to 20.8% (15) in the non-dengue AFI group.

#### Anxiety and Depression in the Overall Study Cohort

In the overall study group, the median HADS anxiety score was 7 (IQR 4-9) and the median HADS depression score was 7 (IQR 4-10). Overall, 80 (41.4%) patients had anxiety and 82 (42.5%) had depression according to the HADS scoring. A total of 110 (56.99%) had either anxiety or depression, while 52 (26.9%) had both anxiety and depression. Among the 80 patients with anxiety in the overall study cohort, 52 (26.9%) had mild anxiety, 20 (10.4%) had moderate anxiety, and 8 (4.2%) had severe anxiety. Among the 82 patients with depression in the overall study cohort, 34 (17.6%) had mild depression, 36 (18.7%) had moderate depression, and 12 (6.2%) had severe depression.

### Unadjusted Analysis of Sociodemographic and Clinical Features Associated with Anxiety or Depression

Among the study cohort, patients with anxiety were more likely to be from lower-income groups than patients from higher-income groups (46.3% vs 31.0, p= .03; Table 2). No sociodemographic features were associated with depression. Anorexia (81.3% vs 67.3%, p=.03), diarrhea (36.3% vs 21.2%, p=.02), and arthralgia/myalgia (81.3% vs 68.1%, p=.04) were associated with anxiety. Anorexia (85.4% vs 64.0%, p=.001), headache (82.9% vs 60.4%, p=.001), vomiting (62.2% vs 41.4%, p=.004), diarrhea (39.0% vs 18.9% p= .002), and abdominal pain (32.9% vs 19.8.0%, p=.04) were associated with depression. Awareness of diagnosis (95.0% vs 86.0%, p= .03) and having lower overall satisfaction scores were associated with anxiety (p=.007). Knowledge and satisfaction scores were not associated with depression.

**Table 2.**
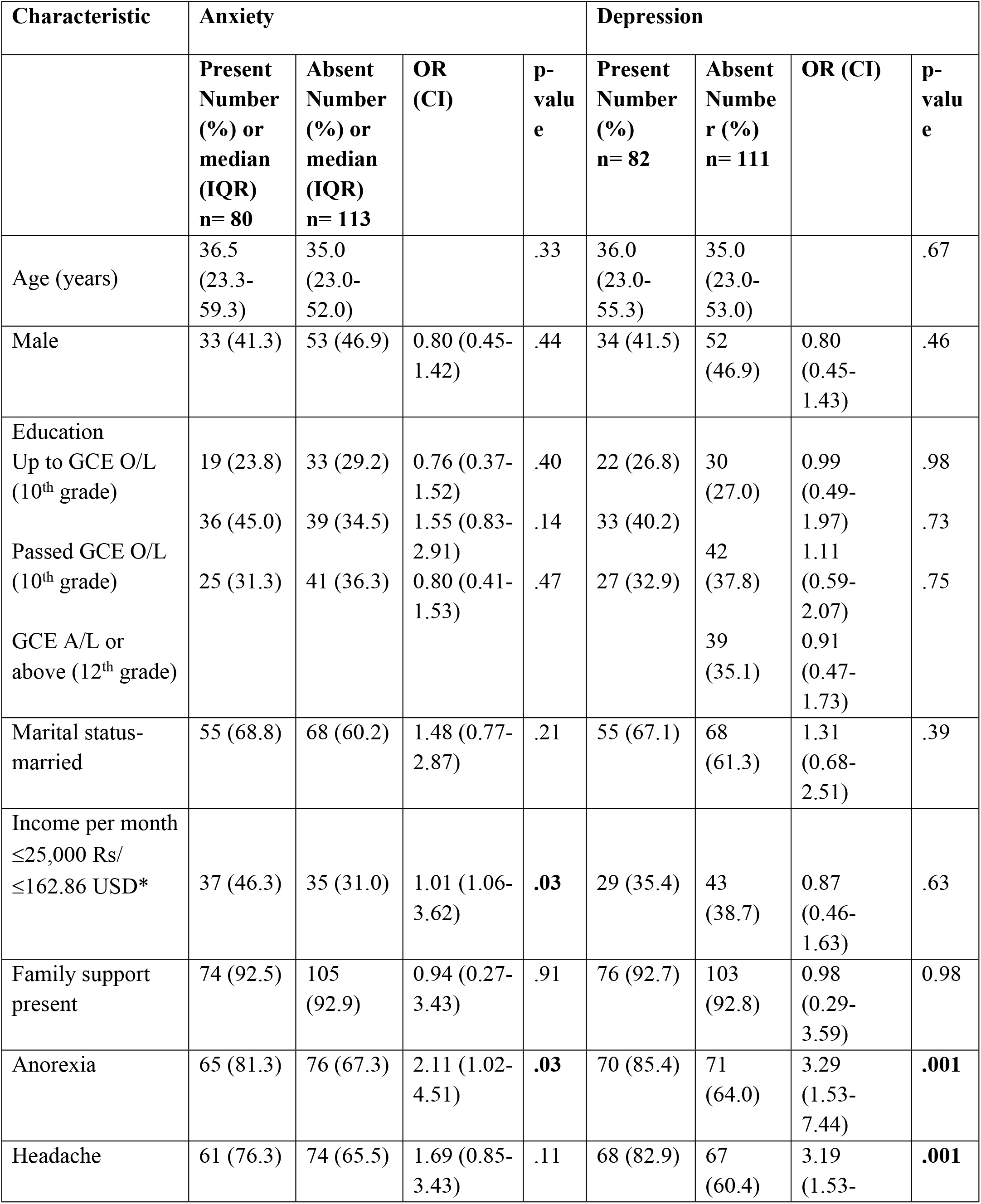
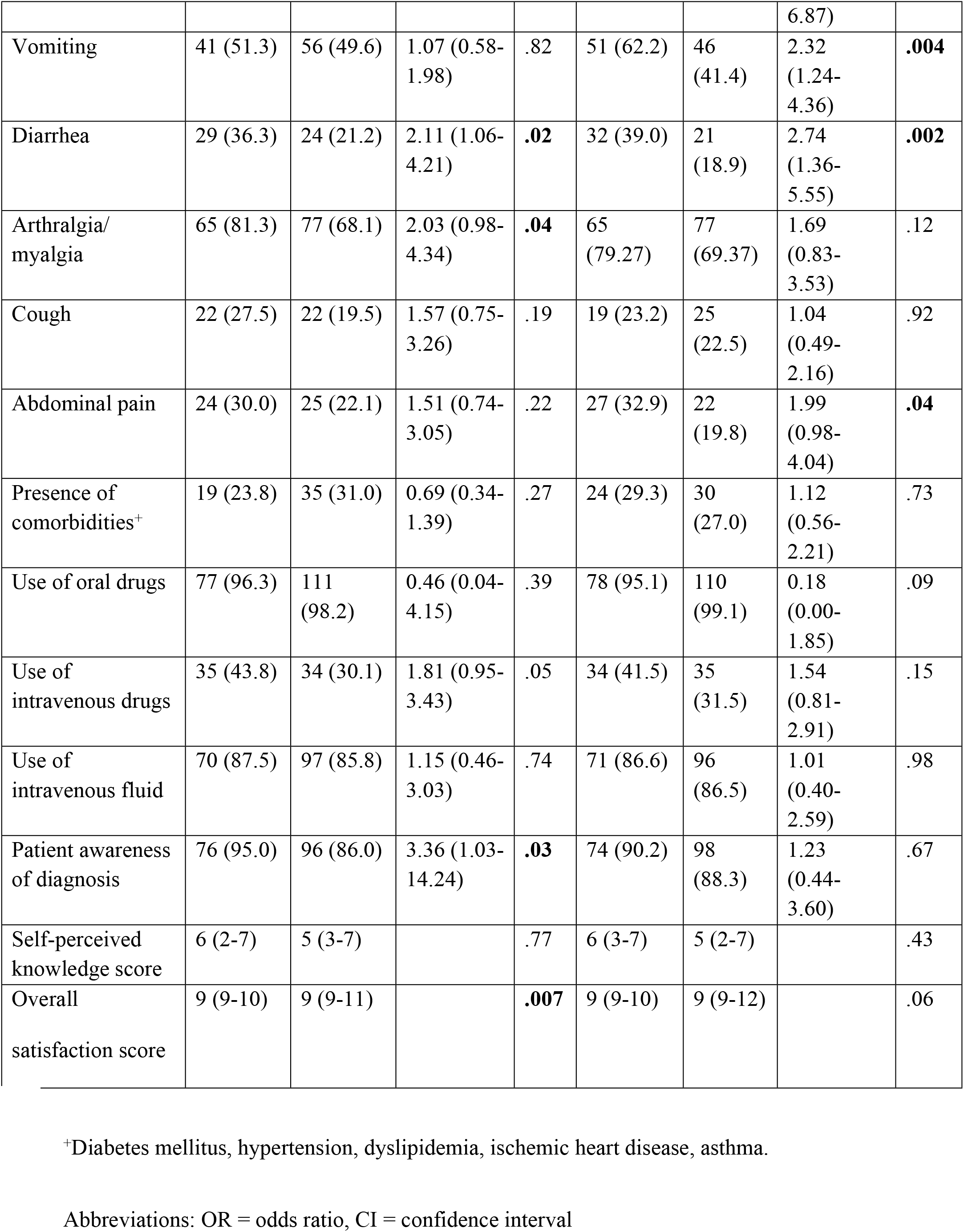
Sociodemographic and clinical characteristics associated with anxiety and depression among patients with acute febrile illness in Sri Lanka.

### Adjusted Analysis of Sociodemographic and Clinical Features Associated with Anxiety or Depression

On multivariable analysis, lower income (1.92, 95% CI 1.03-3.57), patient awareness of their diagnosis (4.30, 1.3-13.76), and overall satisfaction score (OR 0.73, CI 0.58-0.92) were associated with anxiety (Table 3). Anorexia (OR 2.8, CI 1.33-5.87) and diarrhoea (OR 2.29, CI 1.17-4.47) were associated with depression. The presence of family support, presence of comorbidities, or knowledge score regarding current illness were not associated with anxiety or depression on multivariable analysis.

**Table 3.**
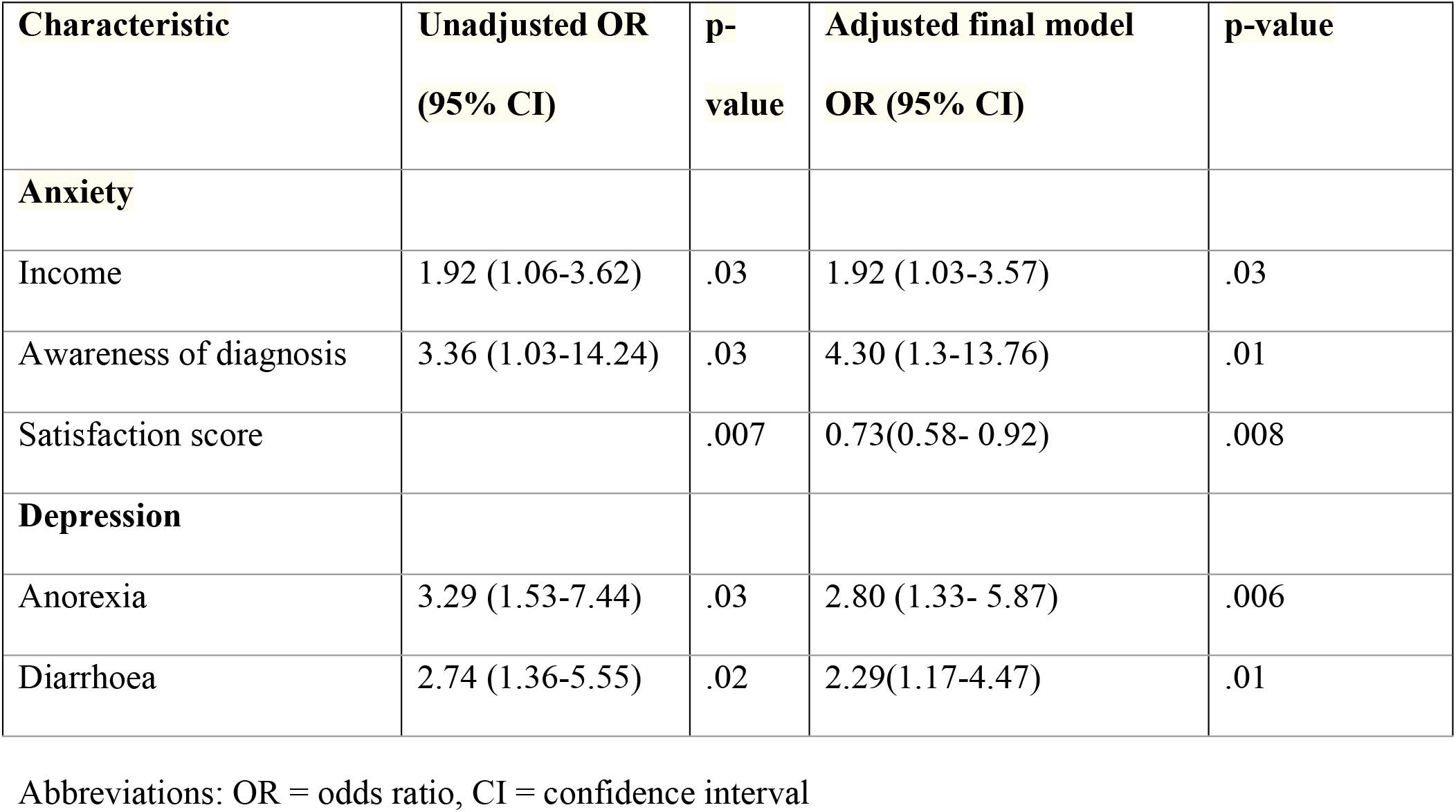
Multivariable analysis of sociodemographic and clinical features associated with anxiety or depression among patients hospitalized with acute febrile illness in Sri Lanka.

| Characteristic | Unadjusted OR<br>(95% CI) | p-value | Adjusted final model<br>OR (95% CI) | p-value |
| --- | --- | --- | --- | --- |
| **Anxiety** |  |  |  |  |
| Income | 1.92 (1.06-3.62) | .03 | 1.92 (1.03-3.57) | .03 |
| Awareness of diagnosis | 3.36 (1.03-14.24) | .03 | 4.30 (1.3-13.76) | .01 |
| Satisfaction score |  | .007 | 0.73(0.58- 0.92) | .008 |
| **Depression** |  |  |  |  |
| Anorexia | 3.29 (1.53-7.44) | .03 | 2.80 (1.33- 5.87) | .006 |
| Diarrhoea | 2.74 (1.36-5.55) | .02 | 2.29(1.17-4.47) | .01 |

Abbreviations: OR = odds ratio, CI = confidence interval

There was no significant difference between dengue versus non-dengue AFI and the presence of anxiety or depression (43.8% vs 37.5%, p= .39; Table 4). No significant difference was also found between the dengue and non-dengue AFI groups with regards to severity of anxiety or depression.

**Table 4.** Anxiety and depression according to the Hospital Anxiety and Depression Scale (HADS) among patients with dengue and non-dengue AFI in Sri Lanka.

|  | **All patients**<br>**Number (%)**<br>**or median**<br>**(IQR)** | **Dengue**<br>**Number (%)**<br>**or median**<br>**(IQR)** | **Non-dengue**<br>**AFI**<br>**Number (%)**<br>**or median**<br>**(IQR)** | **p-value** |
| --- | --- | --- | --- | --- |
| Anxiety present | 80 (41.5) | 53 (43.8) | 27 (37.5) | .39 |
| Depression present | 82 (42.5) | 49 (40.5) | 33 (45.8) | .47 |
| Anxiety and depression<br>both present | 52 (26.9) | 31 (25.6) | 21 (29.2) | .59 |
| Anxiety score | 7 (4-9) | 7 (4-9) | 7 (4-8.5) | .78 |
| Depression score | 7 (4 -10) | 7 (4-10) | 7 (4-11) | .51 |
| Anxiety severity |  |  |  |  |
| Mild | 52 (26.9) | 35 (28.9) | 17 (23.6) | .42 |
| Moderate | 20 (10.4) | 13 (10.7) | 7 (9.7) | .82 |
| Severe | 8 (4.2) | 5 (4.1) | 3 (4.2) | .99 |
| Depression severity |  |  |  |  |
| Mild | 34 (17.6) | 21 (17.4) | 13 (18.1) | .90 |
| Moderate | 36 (18.7) | 21 (17.4) | 15 (20.8) | .55 |
| Severe | 12 (6.2) | 7 (5.8) | 5 (6.9) | .75 |

Abbreviations: AFI = acute febrile illness, IQR = interquartile range

## Discussion

We found a high prevalence of anxiety and depression in patients hospitalized with AFI in Sri Lanka. High prevalence of anxiety and depression was seen in patients with both dengue and non-dengue AFI. A diagnosis of dengue was not specifically associated with a higher prevalence of anxiety or depression when compared to other AFI. The majority of the cohort had mild or moderate levels of anxiety and depression, but a small but significant proportion had severe anxiety or depression. Anxiety was associated with lower socioeconomic status, awareness of illness diagnosis, and lower overall satisfaction score. Depression was associated with gastrointestinal symptoms such as anorexia and diarrhoea. Notably, patients with dengue were more aware of their illness diagnosis and had a higher self-reported knowledge score.

According to our findings, 57% of patients hospitalized with AFI had anxiety or depression as measured by the HADS instrument. No other published studies to-date have assessed anxiety and depression in cohorts with AFI, but several studies have reported similar proportions of anxiety or depression among patients with acute dengue or other specific diseases causing AFI. In a study performed by Hashmi *et al*. using the HADS among patients hospitalized with dengue in Pakistan, 60% met criteria for anxiety and 62.2% met criteria for depression (16). Tran *et al*. showed 64.1% prevalence of anxiety or depression among patients hospitalized with dengue by applying a health-related quality-of-life (HRQOL) survey in Vietnam (5). In Brazil, a case-control study of patients with chikungunya showed significant levels of anxiety and depression (2). In Taiwan, patients with leptospirosis had a 1.58-fold higher risk of depression than the general population (3). During the COVID-19 pandemic, multiple studies evaluated anxiety and depression among patients with COVID-19. In a meta-analysis performed by Deng *et al*., the pooled prevalence of depression and anxiety was 45% and 47%, respectively, among patients with COVID-19 (7). Our study confirms the high levels of anxiety and depression in a cohort with diverse etiologies of AFI, not limited to specific conditions. The underlying reasons for the high prevalence of mental health conditions in patients with AFI are likely multifactorial. Among older adults, cytokine responses to influenza virus have been shown to be associated with mood disturbance, suggesting one possible pathophysiological link (9).

Of note, we found that gastrointestinal symptoms were associated with a high prevalence of depression. A few other studies have reported an association between symptoms related to the gastrointestinal system and anxiety or depression. In Norway, nausea, heartburn, diarrhea, and constipation were associated with anxiety and depression when using the HADS instrument in the general population (17). Among patients at 15 primary care clinics in the US, patients with gastrointestinal symptoms such as stomach pain, constipation and diarrhea were more likely to have anxiety and depression (18). An exact mechanism for the above association is not yet evident.

We found that patients who were aware of their illness diagnosis were more likely to experience anxiety, a finding which contrasts with results from earlier studies. According to a systematic review on the relation between information provision, anxiety, and depression among cancer survivors, satisfied patients and patients with fulfilled information needs had less anxiety and depression (19). Among patients with chronic obstructive pulmonary disease, less disease knowledge was associated with anxiety or depression when using the HADS instrument (20). However, Selinger *et al*. showed that better disease knowledge was associated with greater anxiety among patients with inflammatory bowel disease (21).

In our study, anxiety was higher among patients of a lower socioeconomic status (generally earning less than USD 5 per day). It is likely that the loss of wages due to the acute illness may result in worse mental health for patients. Patients hospitalized with AFI may benefit from coping strategies to deal with the anxiety and depression associated with their acute illness. Strategies such as positive reframing, acceptance of the stressful situation, and use of humor may be helpful interventions in improving the quality of care of these patients.

Some limitations must be noted. Our study was performed at a single center where dengue was prevalent. However, as the causes of AFI were heterogenous, our findings may be generalizable to other settings with AFI. Strengths of this study include the use of a locally validated instrument to measure anxiety and depression and the prospective, systematic collection of data.

## Conclusions

We showed that patients hospitalized with AFI had a high prevalence of anxiety and depression. Gastrointestinal symptoms, lower socioeconomic status, better awareness of illness diagnosis and lower satisfaction score were associated with anxiety or depression. Our findings highlight the need for developing interventions to reduce anxiety and depression among patients hospitalized with AFI in similar settings.

## Data Availability

Data cannot be shared publicly because approval to do so was not originally obtained from the local ethics board. Data may be made available by contacting the Faculty of Medicine, University of Ruhuna Ethical Review Committee, who may grant access on a case-by-base basis for researchers who meet the criteria for access to confidential data.

## Acknowledgments

We would like to acknowledge the patients who were involved in this study.

## Institutional Review Board Statement

The study was conducted according to the guidelines of the Declaration of Helsinki, and approved by the Ethical Review Committee of the Faculty of Medicine, University of Ruhuna. ERC approval No: 14.06.2017:3.10

## Informed Consent Statement

Written informed consent was obtained from all subjects or Parents of children involved in the study.

## Consent for publication

All authors have agreed and give their consented to publish

## Author Contributions

Conceptualization, Champica Bodinayake; Formal analysis, Champica Bodinayake, Wasana Ranaweera Arachchige and L. Gayani Tillekeratne; Investigation, Champica Bodinayake, Wasana Ranaweera Arachchige, Senuri Sahabandu, Aruna Pushpakumara, Piyumi Wijayaratna and Bhagya Jayasekera; Methodology, Champica Bodinayake, Ajith De Silva Nagahawatta, Wasana Ranaweera Arachchige, Senuri Sahabandu, Aruna Pushpakumara, Piyumi Wijayaratna and Bhagya Jayasekera; Project administration, Champica Bodinayake, Ajith De Silva Nagahawatta; Supervision, Champica Bodinayake, Ajith De Silva Nagahawatta; Writing – original draft, Champica Bodinayake; Writing – review & editing, Champica Bodinayake, Ajith De Silva Nagahawatta, Wasana Ranaweera Arachchige, Senuri Sahabandu, Aruna Pushpakumara, Piyumi Wijayaratna, Bhagya Jayasekera and L. Gayani Tillekeratne.

## Data Availability Statement

The datasets generated and/or analysed during the current study are not publicly available due to ethical considerations/ restrictions but may be available from the corresponding author on reasonable request.

## Data Deposition

**Not applicable**

## Geolocation information

**Not applicable**

